# The Public Health Impact of a US Menthol Cigarette Ban on the Non-Hispanic Black Population: A Simulation Study

**DOI:** 10.1101/2022.02.04.22270475

**Authors:** Mona Issabakhsh, Rafael Meza, Yameng Li, Zhe Yuan, Luz Maria Sanchez-Romero, David T. Levy

## Abstract

**Introduction:** As the US Food and Drug Administration (FDA) evaluates implementing a ban on menthol cigarettes and cigars, it is critical to estimate the potential public health effects of such a ban. With high rates of menthol cigarette use by the non-Hispanic Black (NHB) population and important smoking-related health disparity implications, the impact of the ban on this population merits strong consideration.

**Methods:** We apply the previously developed Menthol Smoking and Vaping Model. A Status Quo Scenario is developed using population, smoking and vaping data specific to the NHB population. Estimates from a recent expert elicitation on behavioral impacts of a menthol cigarette ban on the NHB population are used to develop a Menthol Ban Scenario implemented in 2021. The public health impacts of the menthol ban are estimated as the difference between smoking and vaping attributable deaths (SVADs) and life years lost (LYLs) in the Status Quo and the Menthol Ban Scenarios from 2021-2060.

**Results:** Under the Menthol Ban Scenario, overall smoking is projected to decline by 35.1% in 2026, and by 24.0% in 2060 relative to the Status Quo Scenario. With these reductions, SVADs are estimated to fall by about 18.5% and LYL**s** by 22.1%, translating to 255,000 premature deaths averted, and 3.9 million life-years gained over a 40-year period.

**Conclusions:** A menthol cigarette ban will substantially reduce the smoking-associated health impact on the NHB population thereby reducing health disparities.

## INTRODUCTION

Menthol represents 35% of US cigarette sales^1^ and menthol smoking is associated with higher smoking initiation and lower cessation.^2-5^ The US Food and Drug Administration (FDA) has proposed a nationwide ban on menthol cigarettes and cigars,^6^ but will be required to assess its potential public health impact.^7,8^

The Menthol Smoking and Vaping Model (SAVM) estimated the public health impact of a menthol ban on the total US population.^9,10^ However, the model did not explicitly distinguish the impact of the ban on the non-Hispanic Black (NHB) population. Due to their high rates of menthol cigarette use^11,12,13^ and important smoking-related health disparity implications,^14,15^ we apply the previously-developed Menthol SAVM^10^ to evaluate the impact of a menthol cigarette ban on the NHB population.

## METHODS

The SAVM is a publicly available model^16^ which simulates the public health impact of cigarette and nicotine vaping product (NVP) use.^17^ Upon distinguishing menthol and non-menthol cigarette use, menthol SAVM^10^ projects averted deaths and life-years lost (LYLs) from 2013-2016 under Status Quo and Menthol Ban Scenarios. Further model details can be found elsewhere.^10^

### Status Quo Scenario

The NHB observed and projected population and overall mortality rate by single year of age and sex were obtained from CDC Wonder^18,19^ and the US Census Bureau.^20,21^

To initialize the model, menthol and non-menthol NHB smoking prevalence by age and sex are from the 2013/14 Population Assessment of Tobacco and Health (PATH) survey,^22^ with menthol smoking defined as the regular brand flavored to taste like menthol. Current smoking is defined as having smoked ≥100 cigarettes during one’s lifetime and currently smoking at least some days. Smokers become former smokers after having quit for two years, thereby reflecting cessation net of relapse. Regular NVP use is defined in terms of at least 10 days use in the last month.

SAVM^24^ projects never, current, and former smoking prevalence using age-and sex-specific initiation and cessation rates estimated by applying an age-period-cohort model to the NHIS.^25-28^ Using prevalence estimates from the 2014/15 CPS-TUS to calibrate to NHB smoking initiation, we scaled US cigarette initiation rates by 0.91, calculated as the ratio of NHB ages 18-34 to total US ages 18-34 smoking prevalence. To calibrate NHB smoking cessation, we scaled US cessation rates by 0.81, calculated as the ratio of the total US ages 35+ to NHB ages 35+ smoking prevalence.

The proportion of menthol smokers among NHB smokers at age 30 (males 87.8%; females 86.3%), the age at which menthol and non-menthol prevalence rates tended to stabilize, is applied to smoking initiation rates to distinguish menthol and non-menthol smokers. To distinguish NHB menthol vs non-menthol cessation rates, we applied results of a meta-analysis,^29^ which reported that NHB menthol smokers had 12% lower odds of cessation than non-menthol smokers. Similar results were reported by Brouwer et al.^30^

To determine NHB death rates by smoking status, the ratio of NHB to total US population death rates was applied to US never, current, and former smoker death rates.^25,26,31^ Mortality rates of menthol and non-menthol smokers are not distinguished, given limited evidence of differences.^32,33^ To estimate life expectancy for the NHB population, the ratio of 2016 NHB never smokers life expectancy^34^ to 2016 US life expectancy^25,26,31^ was applied to the US life expectancy for the years 2013-2060.

Transitions to NVP use start in 2013. Recent studies^29,35-38^ report lower rates of NVP use among NHB adults than the total adult population. Based on the results from Usidame et al.,^38^ we scaled US NVP prevalence by 70% for the NHB population. Based on lower transition rates from cigarette use to exclusive NVP use among NHB menthol and non-menthol smokers (0.3% and 0.6%) reported by Brouwer et al.,^30^ we estimated NHB menthol smokers switch from smoking to vaping at 50% the rate of non-menthol smokers.

### Menthol Ban Scenario

We model a federal menthol cigarette ban implemented in 2021. An expert elicitation on the impact of a menthol ban^9^ estimated that, of the NHB population who would otherwise initiate into menthol smoking in the absence of a ban, 34.0% would instead become non-menthol smokers, 2.9% illicit menthol smokers, 14.1% NVP users, and 49.0% would not use cigarettes or NVPs. These transitions are applied in the model to the initiation rates of otherwise NHB menthol smokers in 2021 and all future years. Among current NHB menthol smokers ages 18-24, experts expected 9.4% to switch to illicit menthol combustibles, 43.7% to non-menthol combustibles, 23.4% to NVPs and 23.4% to quit all product use.^9^ These transitions are applied to those who were current NHB menthol smokers through age 30 in 2021. Among NHB menthol smokers ages 35-54, experts expected 8.7% to switch to illicit menthol combustibles, 50.9% to non-menthol cigarette use, 15.3% to NVPs, and 25.1% to quit all product use.^9^ These transitions are applied to age 30+ current NHB menthol smokers in 2021. Current non-menthol smokers are unaffected except for those menthol smokers who switch to non-menthol use.

### Outcomes

We estimate the public health impact of a menthol ban as the differences in smoking-and-vaping-attributable deaths (SVADs) and LYLs in the Status Quo and Menthol Ban Scenarios over 2021-2060. Smoking-attributable deaths are estimated as the excess mortality risk for current and former smokers multiplied by their respective populations. Vaping-attributable deaths are estimated assuming 15% of excess smoking risks.^39,40^ Total LYLs are estimated by the number of SVADs multiplied by the expected years of life remaining of a never smoker.

### Validation

We validated pre-ban NHB menthol and non-menthol smoking rate trends against recent evidence from Mattingly et al.,^41^ which was consistent with an earlier study.^23^ Our model projected approximately a 20% decline in NHB adult menthol smoking during the first 5 years (2013-2018) consistent with the 20% decline reported by Mattingly et al.^41^ from 2010-2015. Our 2015 NHB menthol smoking prevalence of 12% is also consistent with Mattingly et al.^41^

## RESULTS

Table 1 shows menthol and non-menthol smoking and NVP prevalence, SVADs and LYLs for NHB adults (ages ≥18). Under the Status Quo, NHB menthol smoking prevalence declines from 12.1% in 2021 to 9.8% in 2026 and 4.4% in 2060, while non-menthol smoking prevalence declines from 2.2% in 2021 to 1.6% in 2026 and 0.6% in 2060. Cumulative SVADs from 2021-2060 of 1,382,385 translate to 17,847,140 LYLs. Under the Menthol Ban, NHB adult menthol smoking prevalence declines from 12.1% in 2021 to 0.7% in 2026 and 0.2% in 2060, while non-menthol smoking prevalence increases from 2.2% in 2021 to 6.7% in 2026 and declines to 3.6% in 2060. Cumulative SVADs of 1,127,270 translate to 13,899,028 LYLs. Comparing the Status Quo and Menthol Ban Scenarios, the model projects 255,115 SVADs and 3,948,112 LYLs averted from 2021-2060.

**Table 1.** NHB Adult Smoking and NVP Prevalence, Smoking and Vaping Attributable Deaths, Life-Years Lost and Public Health Impact, Ages 18 and Abogve, 2021-2060.

| **Status Quo Scenario** |  |  |  |  |  |
| --- | --- | --- | --- | --- | --- |
| **Category** | **Category/Year** | **2021** | **2026** | **2060** | **Cumulative impact \*** |
| **Prevalence** | **Menthol smoker** | 12.1% | 9.8% | 4.4% | -64% |
|  | **Non-menthol smoker** | 2.2% | 1.6% | 0.6% | -73% |
|  | **Total Smokers\*\*** | 14.3% | 11.5% | 5.0% | -65% |
|  | **Former smoker** | 10.5% | 10.5% | 5.5% | -48% |
|  | **Exclusive NVP user\*\*\*** | 2.5% | 3.5% | 6.1% | 144% |
|  | **Former NVP user** | 0.2% | 0.4% | 3.6% | 1700% |
| **Smoking and vaping attributable deaths\*\*\*\*** | **Menthol smoker** | 29,861 | 27,454 | 10,151 | 776,987 |
|  | **Non-menthol smoker** | 8,185 | 6,215 | 1,179 | 138,305 |
|  | **Former smoker** | 8,434 | 9,732 | 9,089 | 412,596 |
|  | **Exclusive NVP user\*\*\*** | 3 | 18 | 938 | 12,486 |
|  | **Former NVP user** | 0 | 0 | 215 | 1,535 |
|  | **Total** | 47,173 | 44,412 | 22,113 | 1,382,385 |
| **Life-years lost** | **Menthol smoker** | 471,851 | 416,832 | 139,218 | 11,318,704 |
|  | **Non-menthol smoker** | 109,219 | 81,674 | 16,468 | 1,833,648 |
|  | **Former smoker** | 83,157 | 93,404 | 84,476 | 3,847,494 |
|  | **Exclusive NVP user** | 119 | 626 | 19,135 | 309,457 |
|  | **Former smoker-NVP user** | 10,906 | 14,873 | 5,131 | 509,637 |
|  | **Former NVP user** | 0 | 0 | 3,420 | 28,200 |
|  | **Total** | 675,251 | 607,410 | 267,849 | 17,847,140 |
| **Menthol Ban Scenario** |  |  |  |  |  |
| **Category** | **Category/Year** | **2021** | **2026** | **2060** | **Cumulative impact \*** |
| **Prevalence** | **Menthol smoker** | 12.1% | 0.7% | 0.2% | -98% |
|  | **Non-menthol smoker** | 2.2% | 6.7% | 3.6% | 64% |
|  | **Total smokers\*\*** | 14.3% | 7.4% | 3.7% | -74% |
|  | **Former smoker** | 10.5% | 12.8% | 5.5% | -48% |
|  | **Exclusive NVP user\*\*\*** | 2.5% | 4.5% | 7.9% | 216% |
|  | **Former NVP user** | 0.2% | 0.5% | 4.6% | 2200% |
| **Smoking and vaping attributable deaths\*\*\*\*** | **Menthol smoker** | 29,861 | 2,427 | 560 | 89,073 |
|  | **Non-menthol smoker** | 8,185 | 20,535 | 5,478 | 494,138 |
|  | **Former smoker** | 8,434 | 10,667 | 9,640 | 451,191 |
|  | **Exclusive NVP user** | 3 | 18 | 1,201 | 16,013 |
|  | **Former smoker-NVP user** | 690 | 2,183 | 884 | 74,951 |
|  | **Former NVP users** | 0 | 0 | 268 | 1,904 |
|  | **Total** | 47,173 | 35,830 | 18,031 | 1,127,270 |
| **Life-years lost** | **Menthol smoker** | 471,851 | 36,081 | 7,580 | 1,286,419 |
|  | **Non-menthol smoker** | 109,219 | 296,779 | 79,254 | 7,000,480 |
|  | **Former smoker** | 83,157 | 104,929 | 88,371 | 4,254,116 |
|  | **Exclusive NVP user** | 119 | 628 | 24,379 | 396,407 |
|  | **Former smoker-NVP user** | 10,906 | 31,350 | 7,853 | 926,363 |
|  | **Former NVP users** | 0 | 0 | 4,290 | 35,242 |
|  | **Total** | 675,251 | 469,767 | 211,729 | 13,899,028 |
| **Public Health Impact: Difference between Menthol Status Quo and Menthol Ban Scenario\*\*\*\*\*** |  |  |  |  |  |
| **Relative Reduction in Prevalence** | **Menthol smoker** | - | -92.9% | -95.5% | - |
|  | **Non-menthol smoker** | - | 318.8% | 500.0% | - |
|  | **Total smokers\*\*** | - | -35.1% | -24.0% | - |
|  | **Total NVP users\*\*\*** | - | 28.6% | 29.5% | - |
| **Gain** | **Averted deaths** | - | 8,582 | 4,082 | 255,115 |
|  | **Averted life-years lost** | - | 137,643 | 56,120 | 3,948,112 |
|  | **% reduction in Averted deaths** | - | 19.2% | 18.5% | 18.5% |
|  | **% reduction in Averted life-years lost** | - | 22.7% | 21.0% | 22.1% |

**Notes:** NVP= nicotine vaping product, Total smokers includes \* The cumulative impact is measured in terms of the relative change from 2021-2060 for prevalence rates (i.e., (2060-2021)/2021) and the sum of the smoking and vaping attributable deaths or life years lost over the years 2021 through 2060. \*\* Total smokers include menthol and non-menthol smokers. \*\*\* Exclusive NVP users includes de novo exclusive NVP users and former smoker now using NVPs. \*\*\*\*\* The number of smoking- and vaping-attributable deaths and life-years lost is rounded to the nearest integer. \*\*\*\*\* The difference between two scenarios includes the comparisons for prevalence in relative terms and for health gains in absolute terms. Relative reduction in prevalence is measured as the relative difference between the Status Quo Scenario and the Menthol Ban Scenario, (i.e. (post ban – pre ban)/pre ban) in year 2026 and 2060; The gain is measured as the increase in the averted deaths and life-years lost from the Status Quo Scenario and the Menthol Ban Scenario, and % reduction in gain is calculated as gain/post ban.

## DISCUSSION

A menthol cigarette ban implemented in 2021 would result in reductions in menthol and non-menthol cigarette use of 35% in 2026 and 24% in 2060. While NVP and non-menthol cigarette use would increase, 255,000 premature deaths would be averted (almost 6,300 per year) and 3.9 million life-years gained (almost 99,000 per year) by 2060.

The ban impact on the NHB population compares favorably to projections for the overall US population.^10^ We found that the ban leads to relative reduction in NHB adult smoking prevalence of 35.1% compared to 14.7% for the US.^10^ The percentage reduction in cumulative NHB averted deaths from 2021-2060 is 18.5% compared to 4.6% for the US,^10^ with a 22.1% relative reduction in NHB cumulative LYLs compared to 7.9% for the US.^10^ Thus, our analysis indicates that a menthol ban would also reduce smoking-related health disparities. Our results are also consistent with earlier modeling results that find disproportionately greater health impacts on the NHB than the general population from a menthol cigarette ban^42^ and past menthol use.^43,44^

Our findings are dependent on the model structure, parameters, and assumptions. While we calibrated the model to smoking and NVP rates, smoking and NVP rates have been subject to recent instability,^45,46^ including in the NHB population.^47,48^ In addition, the model does not distinguish the health impact of exclusive menthol cigarette smokers who switch to cigar use as a result of a menthol ban. For a ban to be effective, especially as it relates to the NHB population, it will be important that it is applied to both menthol cigarettes and flavored cigars, since little cigars are a close substitute for cigarettes.^49-51^ A ban on menthol cigars would yield additional health gains, especially to the NHB population. However, the impact may be reduced if a ban is also imposed on menthol and mint NVP flavored e-cigarettes, since menthol smokers may be less likely to quit smoking in their absence. Finally, the results are also subject to uncertainties regarding impacts of a menthol ban. The effects of a menthol ban on smoking initiation and cessation were based on results of an expert elicitation, and thus dependent on the participating reviewers’ assessments and the process applied in the elicitation.^9^

Our study strongly supports the implementation of a ban on menthol in cigarettes on public health and especially health equity grounds for the NHB population.

## Data Availability

All data produced in the present study are available upon reasonable request to the authors

## Acknowledgments

We would like to thank the Center for the Assessment of Tobacco Regulations (CAsToR) Data Analysis and Dissemination Core for providing data for this work. Research reported in this publication was supported by the National Cancer Institute of the National Institutes of Health (NIH) and FDA Center for Tobacco Products (CTP) under Award Number U54CA229974. The content is solely the responsibility of the authors and does not necessarily represent the official views of the NIH or FDA.

## What This Paper Adds

- The Food and Drug Administration is intending a ban on menthol cigarettes and cigars, and it is critical to show the effect of such a ban on public health. With high rates of menthol cigarette use by the NHB population and important smoking-related health disparity implications, the impact of a menthol ban on NHB individuals merits strong consideration. This study evaluates the public health impacts of a menthol ban on the NHB population.
- The public health implications of a ban on the non-Hispanic Black population have not been considered.
- With a ban on menthol in cigarettes implemented in 2021, NHB adult smoking and vaping attributable deaths are estimated to fall by about 18.5% and years of life lost by 22.1% by 2060, translating to 255,000 premature deaths averted, and 3.9 million life-years gained.
- Our findings strongly support the implementation of a ban on menthol in cigarettes and cigars, resulting simultaneously in considerable health gains and in reductions in health disparities between the NHB and the rest of the US population.

## References

1. Delnevo CD, Giovenco DP, Villanti AC. Assessment of Menthol and Nonmenthol Cigarette Consumption in the US, 2000 to 2018. JAMA Netw Open 2020;3(8):e2013601. doi: 10.1001/jamanetworkopen.2020.13601

2. Malone RE. It’s the 21st century: isn’t it past time to ban menthol cigarette sales? Tob Control 2017;26(4):359–69. doi: 10.1136/tobaccocontrol-2017-053862

3. Wagener TL, Meier E, Hale JJ, et al. Pilot investigation of changes in readiness and confidence to quit smoking after E-cigarette experimentation and 1 week of use. Nicotine Tob Res 2014;16(1):108–14. doi: 10.1093/ntr/ntt138 [published Online First: 2013/10/25]

4. Delnevo CD, Gundersen DA, Hrywna M, et al. Smoking-cessation prevalence among U.S. smokers of menthol versus non-menthol cigarettes. Am J Prev Med 2011;41(4):357–65. doi: 10.1016/j.amepre.2011.06.039 [published Online First: 2011/10/04]

5. Villanti AC, Collins LK, Niaura RS, et al. Menthol cigarettes and the public health standard: a systematic review. BMC Public Health 2017;17(1):983. doi: 10.1186/s12889-017-4987-z [published Online First: 2017/12/30]

6. U.S. Food and Drug Administration. FDA Commits to Evidence-Based Actions Aimed at Saving Lives and Preventing Future Generations of Smokers FDA website2021 [Available from: https://www.fda.gov/news-events/press-announcements/fda-commits-evidence-based-actions-aimed-saving-lives-and-preventing-future-generations-smokers accessed May 6 2021.

7. Schroth KRJ, Villanti AC, Kurti M, et al. Why an FDA Ban on Menthol Is Likely to Survive a Tobacco Industry Lawsuit. Public Health Rep 2019;134(3):300–06. doi: 10.1177/0033354919841011

8. U.S. Department of Health and Human Services FaDA,. Extension of Certain Tobacco Product Compliance Deadlines Related to the Final Deeming Rule: Guidance for Industry Washington DC2017 [Available from: https://www.fda.gov/downloads/TobaccoProducts/Labeling/RulesRegulationsGuidance/UCM557716.pdf accessed November 2 2017.

9. Levy DT, Cadham CJ, Sanchez-Romero LM, et al. An Expert Elicitation on the Effects of a Ban on Menthol Cigarettes and Cigars in the United States. Nicotine Tob Res. 2021 Oct 7;23(11):1911–1920. doi: 10.1093/ntr/ntab121.

10. Levy DT, Meza R, Yuan Z, et al. Public health impact of a US ban on menthol in cigarettes and cigars: a simulation study. Tob Control. 2021 Sep 2:tobaccocontrol-2021-056604. doi: 10.1136/tobaccocontrol-2021-056604.

11. Giovino GA, Villanti AC, Mowery PD, et al. Differential trends in cigarette smoking in the USA: is menthol slowing progress? Tob Control 2015;24(1):28–37. doi: 10.1136/tobaccocontrol-2013-051159

12. Centers for Diseaase Control and Prevention SaTU. Menthol and Cigarettes Atlanta: CDC; 2020 [Available from: https://www.cdc.gov/tobacco/basic_information/tobacco_industry/menthol-cigarettes/index.html accessed November 7 2020.

13. Cornelius ME, Wang TW, Jamal A, et al. Tobacco Product Use Among Adults - United States, 2019. MMWR Morb Mortal Wkly Rep 2020;69(46):1736–42. doi: 10.15585/mmwr.mm6946a4 [published Online First: 2020/11/20]

14. Alexander LA, Trinidad DR, Sakuma KL, et al. Why We Must Continue to Investigate Menthol’s Role in the African American Smoking Paradox. Nicotine Tob Res 2016;18 Suppl 1:S91–101. doi: 10.1093/ntr/ntv209

15. Moolchan ET, Fagan P, Fernander AF, et al. Addressing tobacco-related health disparities. Addiction 2007;102 Suppl 2:30-42. doi: 10.1111/j.1360-0443.2007.01953.x

16. University of Michigan TCORS website. https://tcors.umich.edu/Resources_Download.php?FileType=SAV_Model 2021 [Available from: https://tcors.umich.edu/Resources_Download.php?FileType=SAV_Model.

17. Levy DT, Tam J, Sanchez-Romero LM, et al. Public health implications of vaping in the USA: the smoking and vaping simulation model. Popul Health Metr 2021;19(1):19. doi: 10.1186/s12963-021-00250-7

18. US Population estimates by race, age, and gender 1990-2019 [CDC Wonder], Produced by the U.S. Census Bureau in collaboration with the National Center for Health Statistics (NCHS) [US Population estimates by race, age, and gender 1990-2019 [CDC Wonder], Produced by the U.S. Census Bureau in collaboration with the National Center for Health Statistics (NCHS)]. Available from: https://wonder.cdc.gov/wonder/help/bridged-race.html.

19. Death rate by single age, gender, and race in 1999-2019 [CDC Wonder] [Available from: Underlying Cause of Death, 1999-2019 Request Form (cdc.gov).

20. US Population projections by race, age, and gender 2016-2060 [US Census Bureau] Available from: https://www.census.gov/data/datasets/2017/demo/popproj/2017-popproj.html.

21. Projected Mortality Rates by Nativity, Age, Sex, Race, and Hispanic Origin for the United States: 2017 to 2060 [US Census Bureau] [Available from: 2017 National Population Projections Datasets (census.gov).

22. United States Department of Health and Human Services. National Institutes of Health. National Institute on Drug Abuse, United States Department of Health and Human Services. Food and Drug Administration. Center for Tobacco Products. Population Assessment of Tobacco and Health (PATH) Study Ann Arbor, MI: Inter-university Consortium for Political and Social Research [distributor]; 2018 [Available from: https://www.icpsr.umich.edu/icpsrweb/NAHDAP/studies/36498/datadocumentation.

23. Weinberger AH, Giovenco DP, Zhu J, et al. Racial/ethnic differences in daily, nondaily, and menthol cigarette use and smoking quit ratios in the United States: 2002 to 2016. Preventive medicine 2019;125:32–39.

24. Levy DT, Tam J, Sanchez Romero LM, et al. The Public Health Implications of Vaping in the U.S.: The Smoking and Vaping Model. available as preprint, 2020.

25. Holford TR, Levy DT, Meza R. Comparison of Smoking History Patterns Among African American and White Cohorts in the United States Born 1890 to 1990. Nicotine Tob Res 2016;18 Suppl 1:S16–29. doi: 10.1093/ntr/ntv274

26. Holford TR, Levy DT, McKay LA, et al. Patterns of birth cohort-specific smoking histories, 1965-2009. Am J Prev Med 2014;46(2):e31–7. doi: 10.1016/j.amepre.2013.10.022 [published Online First: 2014/01/21]

27. Jeon J, Holford TR, Levy DT, et al. Smoking and Lung Cancer Mortality in the United States From 2015 to 2065: A Comparative Modeling Approach. Ann Intern Med 2018;169(10):684–93. doi: 10.7326/M18-1250

28. Tam J, Levy DT, Jeon J, et al. Projecting the effects of tobacco control policies in the USA through microsimulation: a study protocol. BMJ Open 2018;8(3):e019169. doi: 10.1136/bmjopen-2017-019169 [published Online First: 2018/03/27]

29. Smith PH, Assefa B, Kainth S, et al. Use of mentholated cigarettes and likelihood of smoking cessation in the United States: a meta-analysis. Nicotine and Tobacco Research 2020;22(3):307–16.

30. Brouwer AF, Jeon J, Cook SF, et al. The Impact of Menthol Cigarette Flavor in the US: Cigarette and ENDS Transitions by Sociodemographic Group. American journal of preventive medicine 2021

31. Holford TR, Meza R, Warner KE, et al. Tobacco control and the reduction in smoking-related premature deaths in the United States, 1964-2012. JAMA 2014;311(2):164–71. doi: 10.1001/jama.2013.285112 [published Online First: 2014/01/09]

32. Hoffman AC. The health effects of menthol cigarettes as compared to non-menthol cigarettes. Tob Induc Dis 2011;9 Suppl 1:S7. doi: 10.1186/1617-9625-9-S1-S7 [published Online First: 2011/06/01]

33. Jones MR, Tellez-Plaza M, Navas-Acien A. Smoking, menthol cigarettes and all-cause, cancer and cardiovascular mortality: evidence from the National Health and Nutrition Examination Survey (NHANES) and a meta-analysis. PLoS One 2013;8(10):e77941. doi: 10.1371/journal.pone.0077941 [published Online First: 2013/11/10]

34. Life expectancy by age, gender, and race in 2013-2017 [CDC National Vital Statistics System] [Available from: https://www.cdc.gov/nchs/nvss/life-expectancy.htm#data.

35. Villanti AC, Mowery PD, Delnevo CD, et al. Changes in the prevalence and correlates of menthol cigarette use in the USA, 2004–2014. Tobacco control 2016;25(Suppl 2):ii14-ii20.

36. Gardiner PS. The African Americanization of menthol cigarette use in the United States. Nicotine & Tobacco Research 2004;6(Suppl_1):S55–S65.

37. Ribisl KM, D’Angelo H, Feld AL, et al. Disparities in tobacco marketing and product availability at the point of sale: results of a national study. Preventive medicine 2017;105:381–88.

38. Usidame B, Hirschtick J, Zavala-Arciniega L, et al. Exclusive and dual menthol/non-menthol cigarette use with ENDS among adults, 2013–2019. Preventive Medicine Reports 2021;24:101566.

39. McNeill A, Brose L, Calder R, et al. Evidence review of ecigarettes and heated tobacco products 2018. A report commissioned by Public Health England. London: Public Health England, 2018.

40. Physicians RCo. Nicotine without smoke. Tobacco harm reduction. London: Royal College of Physicians, 2016.

41. Mattingly DT, Hirschtick JL, Meza R, et al. Trends in prevalence and sociodemographic and geographic patterns of current menthol cigarette use among US adults, 2005–2015. Preventive medicine reports 2020;20:101227.

42. Levy DT, Pearson JL, Villanti AC, et al. Modeling the future effects of a menthol ban on smoking prevalence and smoking-attributable deaths in the United States. American journal of public health 2011;101(7):1236–40.

43. Mendez D, Le TT. Consequences of a match made in hell: the harm caused by menthol smoking to the African American population over 1980–2018. Tobacco control 2021

44. Le TT, Mendez D. An estimation of the harm of menthol cigarettes in the United States from 1980 to 2018. Tobacco Control 2021

45. Gentzke AS, Wang TW, Jamal A, et al. Tobacco Product Use Among Middle and High School Students - United States, 2020. MMWR Morb Mortal Wkly Rep 2020;69(50):1881–88. doi: 10.15585/mmwr.mm6950a1 [published Online First: 2020/12/18]

46. Wang TW, Gentzke AS, Creamer MR, et al. Tobacco Product Use and Associated Factors Among Middle and High School Students - United States, 2019. MMWR Surveill Summ 2019;68(12):1–22. doi: 10.15585/mmwr.ss6812a1 [published Online First: 2019/12/06]

47. Miech R, Leventhal A, Johnston L, et al. Trends in Use and Perceptions of Nicotine Vaping Among US Youth From 2017 to 2020. JAMA pediatrics 2021;175(2):185–90. doi: 10.1001/jamapediatrics.2020.5667 [published Online First: 2020/12/16]

48. Miech RA, Leventhal AM, Johnson LD. Recent, national trends in US adolescent use of menthol and non-menthol cigarettes. Tobacco control 2021 doi: 10.1136/tobaccocontrol-2021-056970 [published Online First: 2021/12/03]

49. Delnevo CD, Giovenco DP, Miller Lo EJ. Changes in the Mass-merchandise Cigar Market since the Tobacco Control Act. Tob Regul Sci 2017;3(2 Suppl 1):S8–S16.

50. Cohn A, Cobb CO, Niaura RS, et al. The Other Combustible Products: Prevalence and Correlates of Little Cigar/Cigarillo Use Among Cigarette Smokers. Nicotine Tob Res 2015;17(12):1473–81. doi: 10.1093/ntr/ntv022

51. Richardson A, Rath J, Ganz O, et al. Primary and dual users of little cigars/cigarillos and large cigars: demographic and tobacco use profiles. Nicotine Tob Res 2013;15(10):1729–36. doi: 10.1093/ntr/ntt053

